# Effect of antihypertensive drugs on glaucoma: a Mendelian randomization and triangulation analysis

**DOI:** 10.1101/2025.01.23.25320990

**Authors:** Yu Huang, Yiliang Liu, Jingze Guo, Yongyi Niu, Hongliang Lin, Jing Liu, Shuilian Chen, Zhuoting Zhu, Yongjie Qin, Xianwen Shang, Denis Plotnikov, Hongyang Zhang, Huan Wang

## Abstract

**Importance:** Glaucoma is one of the world’s leading causes of irreversible blindness and effective interventions with less side effect and less cost were still in need.

**Objective:** To investigate the potential of repurposing antihypertensive drugs to glaucoma treatment and intra ocular pressure (IOP) lowering in cost effective manners.

**Design, Setting and Participants:** Two sample Mendelian Randomization (MR) was used to investigate the genetic effect of antihypertensive drugs on glaucoma protection or IOP lowering. Two strategies (biomarker or eQTL approach) were applied to define the instrumental variable (IV). Genome-wide association study (GWAS) results on systolic blood pressure (SBP) of 526,001 European participants were used as exposure and GWAS on primary open angle glaucoma (POAG, 15,229 cases and 177,473 controls) and on IOP compromising 70832 participants in the UK Biobank were used as the outcome. All analyses were performed between April 2022 and December 2022.

**Exposures:** The SBP lowering effect of the antihypertensive drug targets.

**Main Outcomes and Measures:** POAG or IOP measurements.

**Results:** Using IVs defined by the biomarker approach, genetically-proxied lower SBP through diuretics targets was causally associated with a reduced risk of glaucoma (OR per mmHg reduction in SBP 0.87, 95% CI: 0.97-0.78, p = 0.009 for potassium-sparing diuretics [PSD] and OR=0.86, 95% CI: 0.92-0.81,p =2.67e-05 for loop diuretics [LD]). When using eQTL as IVs, the IVW-MR yielded a causal effect estimate on POAG of 0.88 (95% CI 0.79 -0.97, P = 0.011) per 1 mmHg decrease in blood pressure via PSD targets and 0.92 (95% CI 0.85 – 1, P=0.052) via LD. No consistence effects were observed among different MR methods for other antihypertensive drugs on glaucoma or IOP. After triangulation genetic evidence from MR, epidemiological evidence from observational studies and experimental evidence, the protective effect of PSD/LD on POAG was suggested.

**Conclusions and Relevance:** We found PSD and LD drugs hold promise as potential therapeutic agents to reduce the risk of POAG

**Key Points:** *Question:* Can antihypertensive drugs be repurposed to the treatment of ocular hypertension?

*Findings:* Different Mendelian Randomization (MR) analysis strategies identified that diuretics (potassium-sparing diuretics [PSD) and loop diuretics [LD] might have a protective effect on primary open angle glaucoma (POAG). Furthermore, the triangulation analysis compared the genetic evidence from MR, epidemiological evidence from observational studies and experimental evidence and found a consistent protective effect of PSD/LD on POAG.

*Meaning:* PSD/LD may be repurposed to the treatment of POAG and they may also benefit patients with hypertension as well as POAG.

## Introduction

Glaucoma is one of the world’s leading causes of irreversible blindness and affects an extremely wide range of patients^1^. According to Tham et. al, currently there are 76 million patients suffering from glaucoma worldwide, and this number will reach 100 million by 2040^2^. In terms of treatment, lowering the intraocular pressure (IOP) is the only modifiable risk factor that slows the progression of glaucoma, and current medications are primarily focusing on lowering IOP via different mechanisms^1^. For example, pupil constrictors^3^ and prostaglandins^4^ reduce IOP by increasing the outflow of aqueous humour; while adrenergic receptor blockers^5^, adrenergic receptor agonists^6^, and carbonic anhydrase inhibitors^7^ decrease the production of aqueous humour thus can lower the IOP. However, topical eye drops could induce both topical and systemic side effects in patients, since glaucoma drug targets such as adrenergic, cholinergic, and prostaglandin receptors are widely distributed throughout the body^8^. These side effects include irritation of the eyes^9^, bronchospasm^10,11^ and nausea^12^. Meanwhile, the high cost of certain drugs has discouraged many patients. Therefore, considering the costly drugs and the severe blinding potential of glaucoma, a thorough exploration of the new therapeutic medications for glaucoma is urgently required and may provide patients with better visual quality but less side effects.

Previous epidemiological studies have identified the associations between glaucoma or IOP and blood pressure. Bonomi et. al^13^ found reduced diastolic perfusion pressure is an important risk factor for primary open-angle glaucoma (POAG), and Nigus et. al^14^ found that controlling blood pressure reduces the risk of glaucoma. One of our previous studies explored the causal effect of overall systemic blood pressure (BP) on glaucoma using a Mendelian Randomization (MR) framework^15^. We noted that, although no overall causal relationship can be identified, the effect of BP on glaucoma is heterogeneous, that is, BP may exert different effects on glaucoma or IOP through different mechanisms. Thus, we suspect that lowering BP by different antihypertensive drugs with different pharmacological mechanisms may also lead to different effects on glaucoma.

Antihypertensive drugs can be classified into 12 categories^16^ and previously, only a few trials have investigated the effect of certain types of medication on glaucoma within a limited number of participants^17–20^ and with inconclusive findings. A recent epidemiological study conducted by Yuan et.al^21^ reported that participants who were taking specific antihypertensive medications, such as beta-blocking agents (BBA) and diuretics, were significantly associated with reduced risks of glaucoma onset. This supports our previous hypothesis. However, this observational study may contain ‘spurious associations’ caused by unobserved confounding factors. Furthermore, it didn’t specify which type of diuretic was associated with reduced risks of glaucoma. Mendelian randomization (MR) is an emerging method that can mimick a randomized controlled trial (RCT) and can overcome many of the shortcomings of observational studies^22^. Particularly in recent years, MR has been applied to studies involving drug safety, drug repurposing and drug target discovery^23–25^. In addition, compared with RCTs, this approach provides relatively unbiased estimation with much lower costs. Thus, in the present study, we sought to use two-sample MR to determine the effects of antihypertensive drugs on glaucoma. Comprehensive understanding of antihypertensives and their effect on glaucoma may highlight potentially relevant biological mechanisms for this disease and provide new insight into subsequent basic research and clinical trials of glaucoma drugs.

## Methods

### Overall study design

Figure 1 illustrates the design of this study. We performed a series of two-sample MR analyses to estimate the effects of the antihypertensive drugs on glaucoma and IOP. First, we identified antihypertensive drug classes and their protein targets. Using two different instrument selection strategies (a biomarker and an eQTL approach), SNPs that proxy the protein targets of antihypertensive drugs were chosen as IVs on the basis that they mimicked the action of the drug target^16^. Effect sizes for these SNPs were then extracted from a GWAS meta-analysis of systolic blood pressure (SBP) (226997 participants from the UK Biobank cohort; 299004 participants from the International Consortium for Blood Pressure [ICBP])^15^ to estimate the instrument–exposure association. For the outcome of glaucoma, we estimated the instrument–outcome association using the effect sizes for these same SNPs from a GWAS of glaucoma reported by Gharahkhani et al.^26^. While for the outcome of IOP, we took the SNPs effect size from a GWAS on corneal-compensated intraocular pressure (IOPcc) of 70832 participants in the UK Biobank. For UK Biobank participants, ethical approval was obtained from the National Health Research Ethics Service (Reference 11/NW/0382), and all participants provided (digital) written informed consent. The study adhered to the tenets of the Declaration of Helsinki.

**Figure 1.**
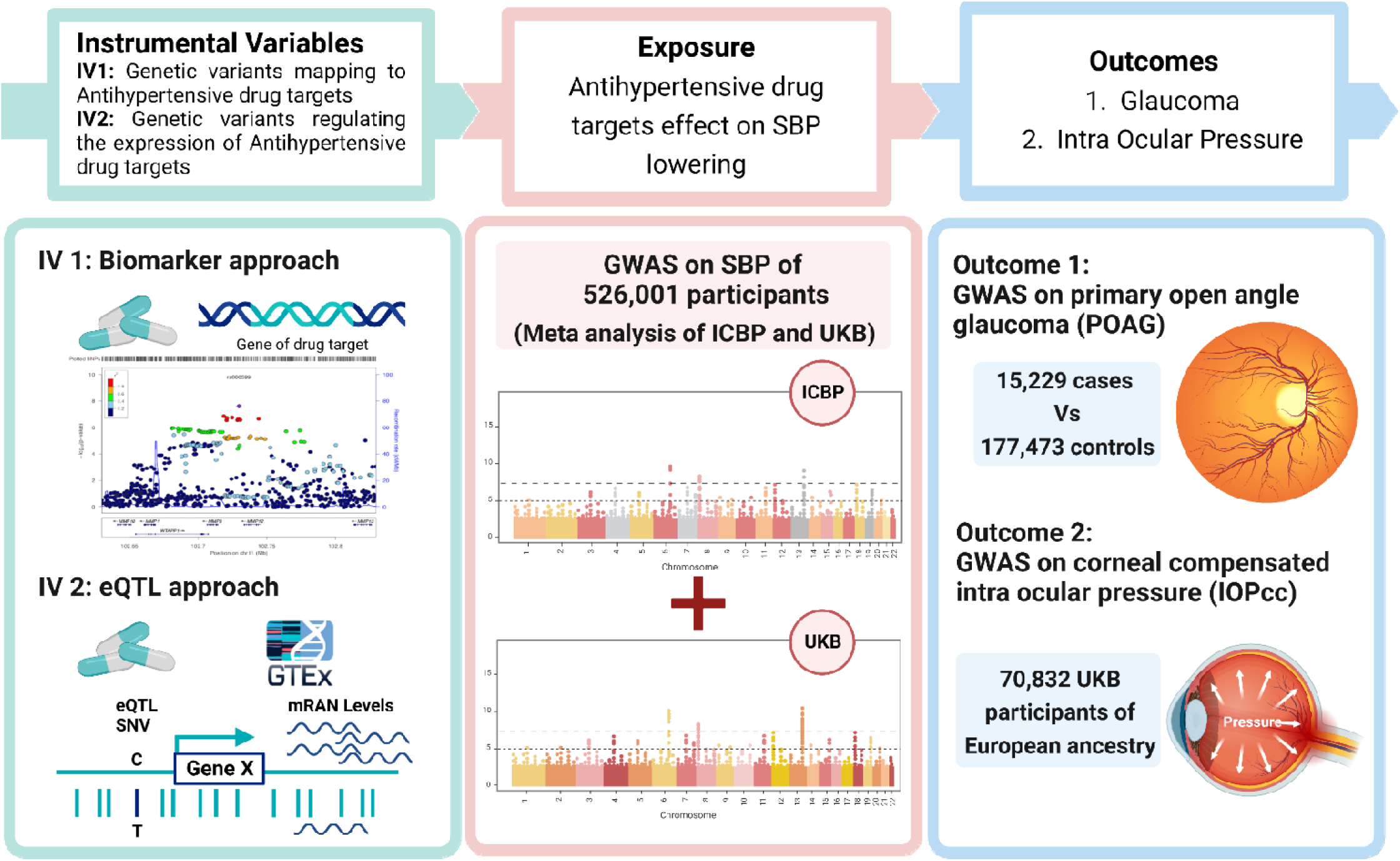
Illustration of the overall study design.

### Systolic blood pressure phenotype

The SBP phenotype was defined using a meta-analysis of two GWASs: one of the UK Biobank cohort and the other of the ICBP cohort^15,27^. The GWAS for SBP from the UK Biobank cohort was performed within 226,997 participants - these participants did not overlap with participants involved in the GWAS for IOPcc. For the SNP quality control, genetic variants with minor allele frequency (MAF) ≥ 1%, missing genotype call rate < 1.5% and Hardy-Weinberg equilibrium p-value < 1.0E-06 were retained in the GWAS. Linear regression was performed to assess each genetic variant and its association with SBP in PLINK 2.0. Covariates including age, sex, genotyping array, body mass index (BMI) and the first 5 ancestry principal components (PCs) calculated by Bycroft et al. al. were adjusted.

A meta-analysis was then conducted to combine summary statistics from the first stage with GWAS for SBP results obtained from International Consortium for Blood Pressure (ICBP). The ICBP study sample had no overlap with UK Biobank and resulted in a meta-analyzed GWAS for SBP of 526001 participants.

### Glaucoma phenotype

Primary open angle glaucoma (POAG) was defined by Gharahkhani et al.^26^. The SNP-POAG regression coefficients were derived from GWAS summary statistics from the first stage of the meta-analysis restricted to people with European ancestry reported by Gharahkhani et al.^26^. After excluding the UK Biobank participants, there were 15,229 cases and 177,473 controls that remained in the GWAS analysis.

### IOP phenotype

In the 70,832 UK Biobank people with European ancestry, a GWAS for IOPcc was conducted. The same criteria of genotype quality control were applied as mentioned above. The first five ancestry PCs, age, sex, genotyping array, and variables were all adjusted in the association study^15^.

### Drug target

Using the British National Formulary^28^, we identified 12 kinds antihypertensive drug classes. They were *adrenergic neurone blocking drugs* (ANB)*; alpha-adrenoceptor blockers* (AAB)*, angiotensin-converting enzyme inhibitors* (ACEI)*, angiotensin-II receptor blockers* (ARB)*, beta-adrenoceptor blockers* (BAB)*, calcium channel blockers* (CCB)*, centrally acting antihypertensive drugs* (CAA)*, loop diuretics* (LD)*, potassium-sparing diuretics* (PSD) *and aldosterone antagonists* (AA), *renin inhibitors* (RI)*, thiazides and related diuretics* (TRD)*, and vasodilator antihypertensives* (VA). As suggested by Walker et.al^16^, via literature search and DrugBank database (https://www.drugbank.ca/; version 5.1.1), we then identified pharmacologically active protein targets of these antihypertension drugs and their corresponding genes. Details of the targets and genes were listed in Supplementary Table 1.

### IV Selection

#### Strategy 1 - a biomarker approach

In strategy 1, according to Dipender et.al^24^ and Maddalena et.al^29^, we selected SNPs that mapped to the encoding regions of the drug targets as IVs for the drug. We first identified the encoding regions of the drug targets by searching the genome build (GRCh37/hg19) information, downloaded from the UCSC Genome Browser (https://genome.ucsc.edu/). To closely resemble the effect of taking an antihypertensive drug, SNPs that fell in the drug target gene region ( ± 200KB) and were associated with SBP (P < 5e-08) were chosen as proxies for the drug targets (except for ANB and PSD where SNPs with P < 1e-05 were used). IVs were further clumped to a pairwise linkage disequilibrium (LD) threshold of r^2^ < 0.4 using the 1000 Genomes European reference panel. Details of the IVs are listed in Supplementary Table 2.

#### Strategy 2 - an eQTL approach

As suggested by Walker et.al^16^ and Zheng et.al^30^, we selected SNPs that regulate the expression of antihypertensive drug targets (eQTLs), as well as have a causal effect on SBP. To identify SNPs that were eQTLs, we identified SNPs to instrument each target using the GTEx project data (Release V8), which contains expression quantitative trait loci (eQTL) analyses of 49 tissues from 838 donors^31^. The most statistically powerful eQTLs (SNP with the smallest nominal p-value for a variant-gene pair defined by GTEx 8) for the drug targets in any tissue were retained for further analysis. To select valid IVs that proxy the SBP-lowering effect of different antihypertensives, we chose SNPs that had a causal effect on SBP. Based on the selected eQTLs, we took their effect on gene expression (derived from GTEX 8) as the effect of exposure together with their effect on SBP (meta-analysis GWAS results) as the effect on the outcome and perform MR analyses for each individual SNP. These MR results were then used to estimate the effect of the drug targets on SBP (i.e. the standard deviation change in SBP per standard deviation change in RNA-expression levels). SNPs with an MR P value less than 0.05 were retained as IVs on the basis that they can 1) regulate the expression level of the drug targets and 2) had a causal effect on SBP. Details of these SNPs were listed in Supplementary Table 3-4.

### Triangulation of genetic and experimental evidence

In aetiological epidemiologys^32^, triangulation of evidence derived from different study design can strengthen the causal inferences. Thus, we triangulated the genetic evidence from MR and epidemiological evidence from observational studies (clinical trials or cohort studies) by searching the literature (from inception to 20 December 2022) to seek different forms of evidence. We searched PubMed from inception to 20 December 2022 for experimental studies about the effects of antihypertensive drugs on glaucoma.

### Statistical analyses

To assess the effect between antihypertensive medications and glaucoma or IOP, a set of two-sample MR analyses were carried out using the R package *‘TwosampleMR’*. For each sets of IVs, the SNP - exposure effect was derived from its effect on SBP and SNP-outcome regression coefficients for these MR analyses were obtained from GWAS for glaucoam^26^ or IOPcc^15^ as mentioned above. Prior to the MR analysis, IVs were further validated by keeping IVs that had strong effect on the exposure (F-statistics > 10) and that were independent by having a pairwise linkage disequilibrium (LD) r^2^ less than 0.1 to nearby SNPs. Inverse-variance weighted (IVW) MR^33^ was used to calculate the causal effect estimates for the main study. As sensitivity analyses for IVW, which assumes no imbalance or horizontal pleiotropy, the effects of the IVs were then calculated using either the MR-Egger^34^, the weighted median^35^, or the mode-based methods^36^. These methods offer a reliable estimate of the causal effect even when some of the IVs violate the MR assumptions. An MR-PRESSO analysis^37^ was conducted as an additional sensitivity analysis for the assumption that all chosen SNPs were valid IVs. The MR-PRESSO study offers an MR causal effect estimate that is intended to be reliable if a proportion of the selected SNPs were invalid IVs due to having strong (outlier) pleiotropic effects on the outcome.

## Results

### Genetic predictors of the antihypertensive drugs targets

Overall, 90 protein targets for 12 antihypertensive drugs were analyzed (Supplementary Table 1). Different SNPs were chosen as instruments for these targets according to the two different MR pipelines. In strategy 1, after mapping the antihypertensive drug regions with SBP GWAS results, 8 drugs (VA, AAB, BBS, ACEIs, TRD, CAA, CCBs and LD) had IVs that reached the genome-wide significant threshold (P < 5e-08 in GWAS for SBP) and 2 drugs (PSD and ANB) had IVs that reached the suggestive significant threshold (5e-08 < P < 1e-05) while no valid IVs were found for renin inhibitors and angiotensin-II receptor antagonists.

In strategy 2, where SNPs with causal effect on regulating the expression of the drug targets (eQTL) were used as instruments, all the drugs had candidate IVs. However, after removing SNPs that had weak effect on the exposure (F statistics less than common threshold of 10), renin inhibitors and adrenergic neuron blockers were not involved for further analysis due to invalid IVs. Details of the IVs for both methods are listed in Supplementary Tables 2-4.

### Effect of antihypertensive drug on glaucoma

Using IVs defined by strategy 1, genetically-proxied lower SBP through PSD targets was causally associated with a reduced risk of glaucoma (OR per mmHg reduction in SBP 0.87, 95% CI: 0.97-0.78, p = 0.009). Additionally, lowering SBP through LD targets also demonstrates a protective effect on glaucoma with an OR=0.86 (95% CI: 0.92-0.81, p =2.67e-05). Sensitivity MR analyses support the effect of lowering SBP through LD targets on glaucoma protection with an estimated effect of OR=0.86 (95% CI: 0.94 -0.79, P =4.87E-04) by weighted median, and limited evidence of pleiotropy (Egger intercept: 0.046, P=0.645) or heterogeneous effects (P=0.736). The effect of lowering SBP through VA targets on glaucoma estimated by IVW was 0.91 (95% CI:1-0.83, P=0.052) but may have been subject to pleiotropy (Egger intercept: 0.053, P=0.039). The weighted median analysis for the effect of lowering SBP through VA targets yielded an estimated effect on glaucoma of 0.88 (95% CI: 0.95-0.82, P =5.28E-04), which was further supported by the result from the MR-PRESSO outlier correction analysis (OR=0.86, P= 0.009) (Figure 2, Supplementary Table 5).

**Figure 2.**
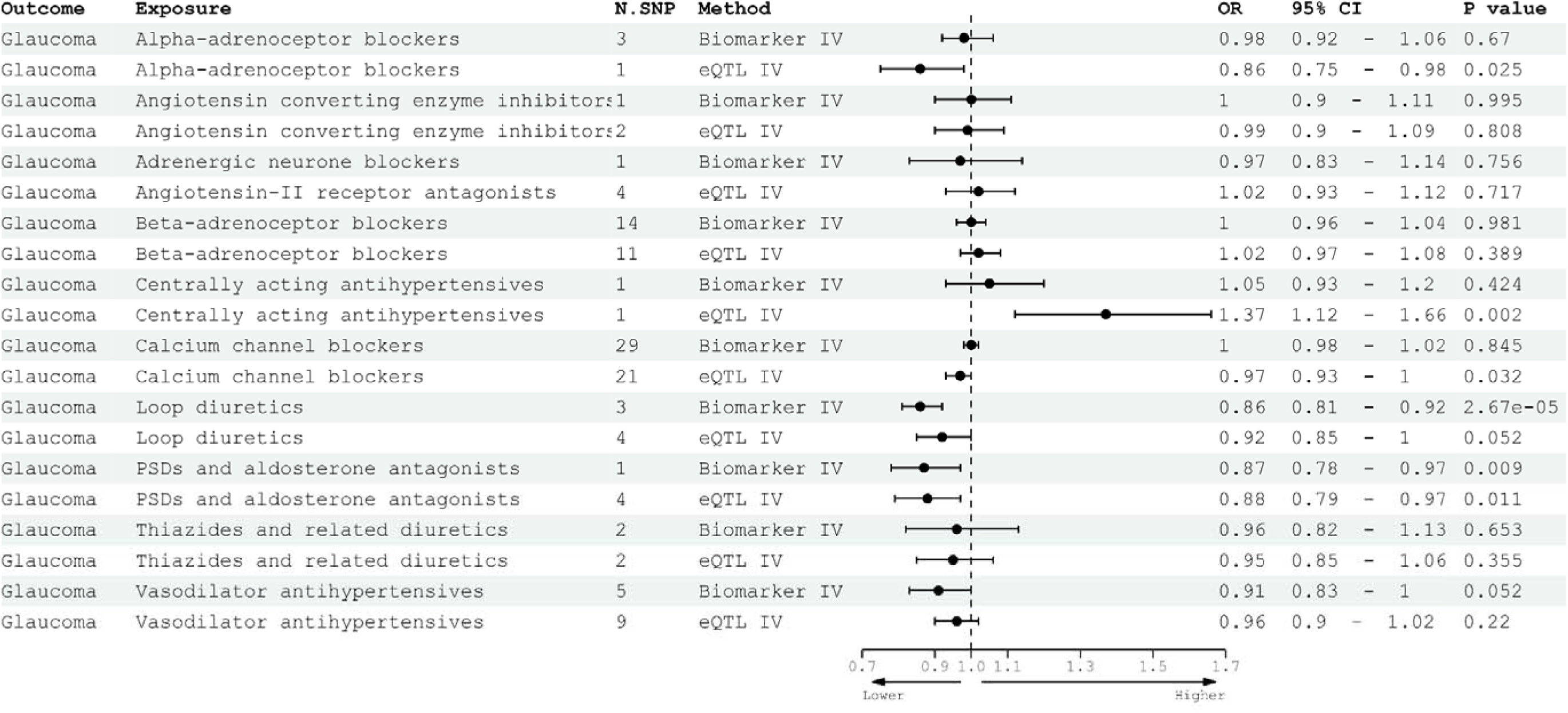
**Association of the antihypertensive drugs with glaucoma in MR analysis**

By using IVs defined by strategy 2, further MR analyses using different sets of IVs were conducted. As for strategy 1, an IVW-MR yielded a causal effect estimate on POAG of 0.88 (95% CI 0.79 -0.97, P = 0.011) per 1 mmHg decrease in blood pressure via PSD targets. Sensitivity analyses were conducted and the weighted median MR supported the main analysis with an effect of OR = 0.85 (95% CI 0.74 - 0.96, P = 0.012). The pleiotropic effect estimated by MR Egger was negligible (Egger intercept: -0.038, P=0.30). The IVW MR causal effect estimate was 0.97 (95% CI: 0.93-1, P = 0.032) for systolic blood pressure lowered by CCB targets, however, it was not supported by the sensitivity analyses. For AAB and CAA, only 1 SNP was used as an IV for each. Lowering BP by these targets yielded a protective effect on glaucoma (OR = 0.86, 95% CI:0.75-0.98, P = 0.025) for the AAB targets but evidence for a potentially harmful effect for the CAA targets (OR = 1.37, 95% CI:1.12-1.66, P = 0.002). (Figure 2, Supplementary Table 6).

### Effect of antihypertensive drug on IOP

The effect of antihypertensive drugs on IOP was also evaluated by MR using the two instrument selection strategies. In strategy 1, an IVW MR analysis estimated the causal effect of lowering systolic blood pressure via VA targets on IOP to be -0.17 mmHg IOPcc per 1 mmHg SBP reduction by VA (95% CI: -0.3 to -0.03; P = 0.016). In the sensitivity analysis, weighted median MR yielded a causal estimate of -0.15 mmHg IOPcc per 1 mmHg SBP reduction (95%CI: -0.25 to -0.06; P = 0.001), however, after correction for two outlier SNPs, the effect in the MR-PRESSO analysis attenuated (P = 0.13). It was noticed that for AAB and CCB, reducing SBP by these drugs might cause an increase in IOP, with a causal effect estimate from the IVW of +0.09 mmHg (95% CI 0.01 to 0.18; P = 0.032) and +0.03 mmHg (95% CI 0.01 to 0.06; P = 0.005) respectively. The causal effect estimates using the weighted median approach were +0.1 mmHg (95% CI 0.01 to 0.19; p = 0.032) and +0.03 mmHg (95% CI -0.01 to 0.06; p = 0.136) respectively. (Figure 3, Supplementary Table 7).

**Figure 3.**
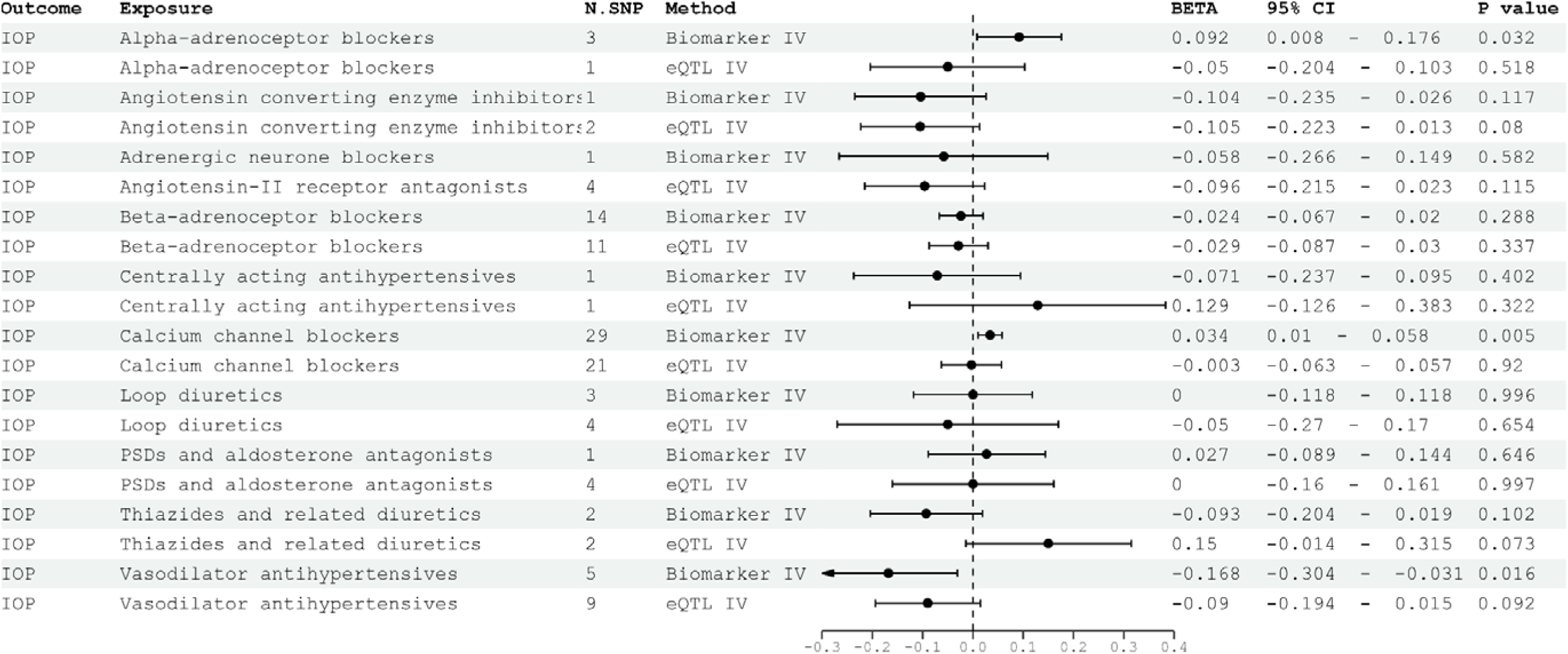
**Association of 12 antihypertensive drugs with IOP in MR analysis**

After performing the MR analysis using IVs selected in strategy 2, the IVW and sensitivity analysis provided limited evidence for a causal relationship between antihypertensive drug targets mentioned above and IOPcc. Heterogeneous effects were detected in the analyses of CCB, LA, VA, and PSD targets. Correcting for outlier SNPs by MR-PRESSO did not impact this inference. (Figure3, Supplementary Table 8).

### Triangulation of genetic and literature evidence

To overcome different inherent biases in different study designs, we triangulated the existing evidence from the literature with the genetic evidence we obtained from this study (Table 1). Collectively, the evidence indicates that diuretics, especially targets of PSD and LD, may exert a protective effect on glaucoma with similar effect size estimated by MR and epidemiological study. The effect of CCB targets on glaucoma was contradictory. Interestingly, both genetic and epidemiological evidence suggested that CAA targets may even increase the risk of glaucoma. Meanwhile, consistent with previous findings, we found a protective effect of AAB targets on glaucoma.

**Table 1.** Triangulation of genetic and literature evidence.

| Drug | Strategy 1<br>IVW<br>OR (95%CI) | Strategy 1<br>Sensitivity<br>OR (95%CI) | Strategy 2<br>IVW<br>OR (95%CI) | Strategy2<br>Sensitivity<br>OR (95%CI) | Literature<br>OR (95%CI) | Experimental evidence |
| --- | --- | --- | --- | --- | --- | --- |
| PSD | 0.87(0.78 - 0.97) | NA | 0.87( 0.79 - 0.97) | 0.85(0.74 - 0.96) | 0.99(0.82 - 1.19)<sup>46</sup><br>0.85(0.77–0.95)<sup>21</sup> | PSD might directly protect loss of retinal ganglion cells (RGCs) and optic nerve fibers independent of IOP<sup>41</sup>. |
| LD | 0.86(0.81 - 0.92) | 0.86(0.79 - 0.94) | 0.92(0.85 - 1) | 0.92(0.82 - 1.02) | 0.93(0.85 - 1.03)<sup>46</sup><br>0.9 (0.5 - 1.9)<sup>20</sup><br>0.85 (0.77–0.95)<sup>21</sup> | LD can reduce IOP in rabbit and monkey<sup>47</sup>. |
| CCB | 1(0.98 - 1.02) | 1(0.97 - 1.03) | 0.97(0.93 - 1) | 0.98(0.93 - 1.02) | 1.8 (1.04 - 3.2)<sup>20</sup><br>1.26 (1.18 - 1.35)<sup>46</sup><br>1.00 (0.92–1.09)<sup>21</sup> | Inconsistent effect on IOP in animal models <sup>48-50</sup> |
| CAA | 1.05 (0.93 - 1.2) | NA | 1.37(1.12 - 1.66) | NA | 1.28 (1.08 - 1.51)<sup>46</sup> | CAA can reduce the IOP in animal models<sup>51</sup> |
| AAB | 0.98 (0.92 - 1.06) | 0.99 (0.9 - 1.07) | 0.86 (0.75 - 0.98) | NA | 0.91 (0.78 - 1.05)<sup>46</sup> | Topical applicaiton of AAB had a protective effect on glaucoma.<sup>52,53</sup> |

## Discussion

In the present study, we investigated the effect of different antihypertensive mechanisms on glaucoma using an MR approach. We selected BP-lowering genetic variants by 2 strategies as proxies for the effects of antihypertensive drug classes. Using different IVs, we found lowering BP through PSD, CCB and LD might be associated with lower risk of glaucoma, which will provide novel directions for the drug repurposing for glaucoma.

Drug repurposing studies using MR approaches intend to identify novel applications of existing drugs in a more rapid and cost-effective manner compared to traditional drug development. Previously, numerous epidemiological studies have suggested that antihypertension drugs may influence glaucoma, motivating us to investigate their potential treatment effect on glaucoma using MR. For instance, in an epidemiological study that included all Danes aged 40-95 years from 1996-2012, Anna et al. found that taking antihypertensive drugs significantly reduced the risk of glaucoma^38^. Moreover, Yuan et al.^21^ further investigated the relationship between taking different anti-hypertensive drugs and the risk of developing glaucoma using a cross-sectional study, and their study found that taking BBS and diuretics protected against glaucoma, similar to our findings. Furthermore, the present study provides additional evidence for specific diuretic drugs and CCB targets as potential repurposing opportunities for glaucoma treatment.

As for diuretic drugs, previous explorations of the mechanisms by which diuretics reduce the risk of glaucoma have provided both basic and clinical evidence for our study. Lama et al.^39^ found that in isolated human eyes, bumetanide increased atrial outflow by inhibiting the Na-K-2Cl transporter and thus may have an effect on IOP. Through intraocular injection of Ethacrynic (a PSD) in 5 patients with glaucoma, Shlomo et al.^40^ found that the IOP of all patients decreased significantly after intraocular injection, and there were no obvious side effects. An in vitro study has also demonstrated the neuroprotective effect of PSD targets: Nitta et al.^41^ found that spironolactone antagonized aldosterone directly protected RGC cells in mice treated with aldosterone. However, these studies are either limited to animal experiments, unconvincing due to small sample sizes, or lacking in explanation for the specific mechanism. Compared with these studies, our data were derived from real-word human studies with large sample sizes.

As for CCBs, the mechanism by which CCB drugs reduce intraocular pressure is unclear. Previous studies have put forward many different hypotheses: one hypothesis is that there is a gap junction regulated by calcium in the ciliary epithelium. CCB drugs affect aqueous humor production by regulating this gap junction, and CCB drugs may change aqueous humor production by interfering with the gap junction of the ciliary body^42,43^. Another is that CCB drugs affect aqueous humor outflow by inhibiting the contraction of trabecular meshwork cells^44^. In addition, other studies have found that CCB drugs may be involved in activating the matrix remodeling of the optic nerve head, thereby slowing the progression of glaucoma^45^. Our study found that CCB targets are associated with a reduced risk of glaucoma but increase IOP. It may be necessary to further explore the mechanism of CCBs in delaying the progression of glaucoma to fully evaluate their safety and effectiveness.

Besides drug repurposing, the present study also provides an insight into potential mechanisms. In the existing clinical guidelines, IOP is the only known risk factor affecting the clinical progression of glaucoma, so most current clinical drugs are aimed at reducing IOP. While previous studies have found that some drugs may delay the progression of glaucoma through mechanisms other than lowering intraocular pressure, such as the neuroprotective effects of PSD and CCBs^41,45^, our study found that some drugs are related to reducing the incidence of glaucoma, but not related to the reduction of intraocular pressure, such as CCB targets. This may suggest that there are other ways to delay the clinical progression of glaucoma. These novel findings may be relevant to the development of new drugs for glaucoma and our understanding of the pathophysiological mechanisms of glaucoma.

Despite using MR to minimize confounding and consistency with RCTs, this study has several limitations. First, MR relies on three assumptions, i.e., the genetic instruments are associated with the exposure, are not associated with potential confounders, and the association of the genetic instruments with the outcome is exclusively through the exposure. To satisfy the first of these assumptions, we used two strategies to select SNPs related to the expression of genes or encoding the genes regulating the relevant antihypertensive target proteins. Secondly, the data used in this study are from European populations, so the applicability of the findings of this study to other populations worldwide is not entirely clear. The outcome data in this study were all from POAG patients and do not indicate the effect of antihypertensive drug targets on other types of glaucoma. Thirdly, it was not possible to instrument all antihypertensive drug targets, despite the use of multiple instrument selection strategies in our study. Finally, this study cannot explore effects of antihypertensive drug targets on glaucoma that do not act through SBP.

## Conclusions

In this study, we selected BP-lowering variants by two strategies to proxy the effect of antihypertensive drug targets and investigated their effect on glaucoma in Mendelian randomization analyses. For both strategies, we found lowering blood pressure though PSD and LD targets was associated with a lower risk of glaucoma. This provides new evidence for research into glaucoma treatments.

## Data Availability

All data produced in the present study are available upon reasonable request to the authors.

## Acknowledgement

This research was conducted using the UK Biobank resource. We thank the participants of the UK Biobank.

## Author contributions

Study concept and design: Wang H, Zhang HY, Plotnikov D, Huang Y; Access to raw data: Huang Y, Liu YL, Guo JZ, Wang H, Plotnikov D; Acquisition, analysis, or interpretation: All authors; Drafting of the manuscript: Liu YL, Huang Y, Wang H, Guo JZ, Niu YY; Critical revision of the manuscript for important intellectual content: Zhang HY, Wang H, Shang XW, Lin HL, Qin YJ; Statistical analysis: Huang Y, Liu YL, Chen SL; Obtained funding: Wang H, Huang Y, Zhang HY; Administrative, technical, or material support: Liu J, Chen SL; Study supervision: Wang H, Plotnikov D, Zhang HY.

## Data Availability

All data produced in the present study are available upon reasonable request to the authors.

